# Prevalence and factors associated with institutional delivery service utilization among mothers in government health facilities of Tamankhola Rural Municipality: A community-based cross-sectional study

**DOI:** 10.1101/2025.07.14.25331548

**Authors:** Sandip Dhungana, Shirish Sharma, Archana Shah, Karuna Bhandari, Ganesh Baruwal, Ashok Bhurtyal

## Abstract

**Introduction:** Despite constitutional and legal guarantees for safe motherhood and reproductive health in Nepal, significant disparities persist in institutional delivery service utilization, particularly among marginalized groups and rural regions. According to the Nepal Demographic and Health Survey (NDHS) 2022, 19% of births still occur at home. This study assessed the prevalence and determinants of institutional delivery among mothers in government health facilities of Tamankhola Rural Municipality to address this gap.

**Methods:** A descriptive cross-sectional study was conducted from January to June 2022 in Tamankhola Rural Municipality, Baglung. Using simple random sampling from immunization records, 126 mothers of children born between 2077/11/01 and 2078/11/01 B.S. were interviewed with a pretested structured questionnaire. Descriptive statistics and logistic regression analyses identified factors associated with institutional delivery.

**Result:** The mean age of mothers was 25.27 years; 68.25% were Dalit and 69.05% belonged to lower socio-economic status. The prevalence of institutional delivery was 90.48%. Most mothers (89.68%) had four or more Ante-Natal Care (ANC) visits, and 98.41% received counseling, primarily on institutional delivery while only 5.56% of the mothers received counseling on Post-Natal Care (PNC). While 88.09% had knowledge of birth preparedness, only 46.82% recognized danger signs of pregnancy. A statistically significant association was found between birth order and institutional delivery (p=0.04), with mothers having more than one child being significantly less likely to utilize institutional delivery services (AOR=0.11; 95% CI: 0.01–0.97).

**Conclusion:** The prevalence of institutional delivery was high and significantly associated with birth order of the child. Counseling on Post-Natal Care and information on danger signs during pregnancy. Despite the high institutional delivery rate, 9.52% still delivered at home, mainly due to poor road conditions. Strengthening PNC counseling, improving health education on danger signs during pregnancy, and addressing structural barriers are essential to address the problem of home delivery in Tamankhola rural municipality.

## Introduction

Article 38 of the Constitution of Nepal 2072 B.S. guarantees every woman safe motherhood and reproductive health rights (1). Safe Motherhood and Reproductive Health Act, 2018 also guarantees the right of every woman to receive antenatal care service, obstetric care, and emergency obstetric and newborn care(2). Government of Nepal (GoN) encourages every woman to have institutional delivery through “Safe Motherhood Programme” where women receive transportation incentives as per their ecological residence i.e., Rs.1000 in Terai, Rs.2000 in hills, and Rs.3000 in Mountains to reduce maternal and neonatal morbidity and mortality in line with Sustainable Development Goal (SDG)(3).

To reach the SDG target of Maternal Mortality Ratio (MMR) to less than 70 per 100,000 live births and Neonatal Mortality Rate (NMR) to less than 12 per 1000 live births, delivery of women in a health institution by the skilled birth attendant is a prime strategy(4). According to Nepal Demographic and Health Survey (NDHS) 2022, there are still 19% of home delivery(5) and as per NDHS 2016, there are still 57% of home delivery in the mountain region, 37.4% in the hilly region, and 39.2% in the Terai region(6).

Tamankhola Rural Municipality is one of the 10 local levels in Baglung district within Gandaki Province, which is formed by merging the three former Village Development Committees (VDCs) of Bongadobhan, Taman, and Khungkhani into a single rural municipality with six wards and 740 households(7). There was disparity among home delivery, with 3.3% of home delivery in urban areas and 23.3% of home delivery in rural areas in Gandaki province as per NDHS 2022(5). There were still 161 MMR per 100,000 live births in Gandaki province, and 39% of the maternal deaths were in the hilly region as per the National Population and Housing Census (NPHC) report 2021(8). Though Gandaki province has lowest neonatal mortality and under-five mortality across all the provinces, there was huge disparity among the urban and rural areas in the province with two neonatal deaths per 1000 live births in urban area, 20 neonatal deaths per 1000 live births in rural area, seven under-five mortality per 1000 live births in urban area and 52 under-five mortality per 1000 live births in rural area(5)

The trends in the percentage of women giving birth in health facilities were lowest for Dalit and Janajati ethnic groups as per NDHS 2006, NDHS 2011, and NDHS 2016(6). Similarly, a study conducted by Chaurasiya et.al (2019) to investigate the rate of and factors associated with the institutional delivery among the Dalit women of the Mahottari, Nepal found that only 30% of the mothers had institutional delivery(9). The residents of Tamankhola Rural Municipality are mostly Dalit (56.73%) and Janajati (33.24%) as per the Annual Development Plan for Fiscal Year (FY) 2081/082 B.S.(10). No research has been done yet to assess the prevalence and factors associated with institutional delivery in this rural municipality. This research was done to fill the research gap by assessing the prevalence of institutional delivery and factors associated with institutional delivery service utilization among mothers in government health facilities of Tamankhola Rural Municipality.

## Methods

### Study design

A descriptive cross-sectional study design was used to assess the prevalence of institutional delivery and factors associated with institutional delivery service utilization among mothers in Tamankhola Rural Municipality.

### Study duration

The total duration of the study was six months from January 2022 to June 2022, and the field-based data collection was done for 20 days between 15^th^ April 2022 to 5^th^ May 2022.

### Sampling frame

All the focal person of government health institutions of Tamankhola Rural Municipality were approached to get an updated and complete list of children born and immunized during the period 2077/11/01 B.S. to 2078/11/01 B.S. from the immunization register as the rural municipality has 100 % BCG coverage and the sampling frame included 185 mothers who were the study population of the research study.

### Sample size and sampling technique

Based on the prevalence of institutional delivery in Gandaki province as per NDHS 2016 i.e., 0.683(6) using Cochran’s sample size formula, 120 was the minimum sample size, and taking a 10% non-response rate from a similar study conducted by Shah et.al (2015) in six VDCs of Chitwan district of Nepal to find out the factors affecting institutional delivery (11), the final sample size was 132. A simple random sampling method (fishbowl method) was applied as a sampling method for the identification of respondents from the sampling frame for data collection.

### Inclusion and exclusion criteria

Mother of children born between 2077/11/01 B.S.to 2078/11/01 B.S. and immunized in any health institution providing immunization service of Tamankhola Rural Municipality and who provided written consent to participate in the research study were included in the study while the mother who were mute and who had migrated from the Tamankhola rural municipality during the period of data collection were excluded from the research study.

### Validity of the research tool

Research instruments were prepared based on an extensive literature review, pre-testing among 10% of the sample size in Bobang rural community of Dhorpatan municipality, Baglung district, and modified in consultation with the research supervisor to ensure content validity(12).

### Ethical consideration

Before data collection, approval from the health section of Tamankhola Rural Municipality was taken by submitting a letter from the Institutional Review Committee (IRC), Institute of Medicine (IoM) [Ref:389(6-11)(2078/079)]. Written informed consent was taken from each participant before the data collection. Anonymity and confidentiality of the study participants were maintained by giving them unique codes.

### Data collection and analysis

Data was collected over 20 days using a structured questionnaire through face-to-face interviews, and a total of 126 quantitative data were collected. The research study had a 95.45% response rate.

Quantitative information was entered in Epi-data version 3.1, then was transferred to MS Excel for cleaning and development of suitable codes based on the literature review. Data from MS Excel was imported into EZR for descriptive and inferential statistics. The descriptive statistics were presented using frequency, percentage, mean/median, and standard deviation/interquartile range. Kolmogorov-Smirnov Test, which was used for more than 50 samples, was used for testing the normality since the total samples collected were 126(13). Bivariate analysis of dependent and independent variables was carried out. Variables having a p-value less than 0.25 in bivariate analysis were taken into consideration for multivariate logistic regression(14). Multicollinearity was assessed using the Variance Inflation Factor (VIF), and variables with VIF ≥2 were sequentially dropped, retaining only those variables with VIF < 2 for multivariate logistic regression analysis(14).

## Results

The mean age of the mothers was 25.27 with a Standard Deviation (SD) of 5.41. The majority (68.25%) of the mothers were of Dalit ethnicity. More than half (58.73%) of the mothers had ≥6 family members. Similarly, more than half (53.17%) of the mothers were living in a joint family. More than half (59.52%) of the mothers and the majority (60.32%) of their husbands had a basic and below level of education. Almost all (95.24%) of the mothers and more than half (53.97%) of their husbands were involved in household work or agriculture. The majority (69.05%) of the respondents had lower Socio-economic Status (SES) (Table 1).

**Table 1:** Socio-demographic characteristics of mothers.

| **Variables** | **Frequency (n)=126** | **Percentage (%)** |
| --- | --- | --- |
| **Age** |  |  |
| <20 | 14 | 11.11 |
| ≥20 | 112 | 88.89 |
| Mean ± SD (min, max) | 25.27±5.41 (17,45) |  |
| **Ethnicity** |  |  |
| Dalit | 86 | 68.25 |
| Others | 40 | 31.75 |
| **Family size** |  |  |
| <6 | 52 | 41.27 |
| ≥6 | 74 | 58.73 |
| **Family type** |  |  |
| Nuclear Family | 45 | 35.71 |
| Joint Family | 67 | 53.17 |
| Extended Family | 14 | 11.11 |
| **Educational status** |  |  |
| Basic education and below | 75 | 59.52 |
| Secondary education and above | 51 | 40.48 |
| **Educational status of the husband** |  |  |
| Basic education and below | 76 | 60.32 |
| Secondary education and above | 50 | 39.68 |
| **Occupational status** |  |  |
| Household work or agriculture | 120 | 95.24 |
| Others | 6 | 4.76 |
| **Occupation of husband** |  |  |
| Household work or agriculture | 68 | 53.97 |
| Others | 58 | 46.03 |
| **Socio-economic Status (SES)** |  |  |
| Lower SES | 87 | 69.05 |
| Middle SES | 39 | 30.95 |

Most (89.68%) of the mothers had four or more Ante-Natal Care (ANC) visits, and almost all (98.41%) of the mothers received counselling during their ANC visit from the health workers. Regarding the components of ANC counselling, almost all (96.03%) mothers received counselling for having an institutional delivery (Table 2).

**Table 2:**
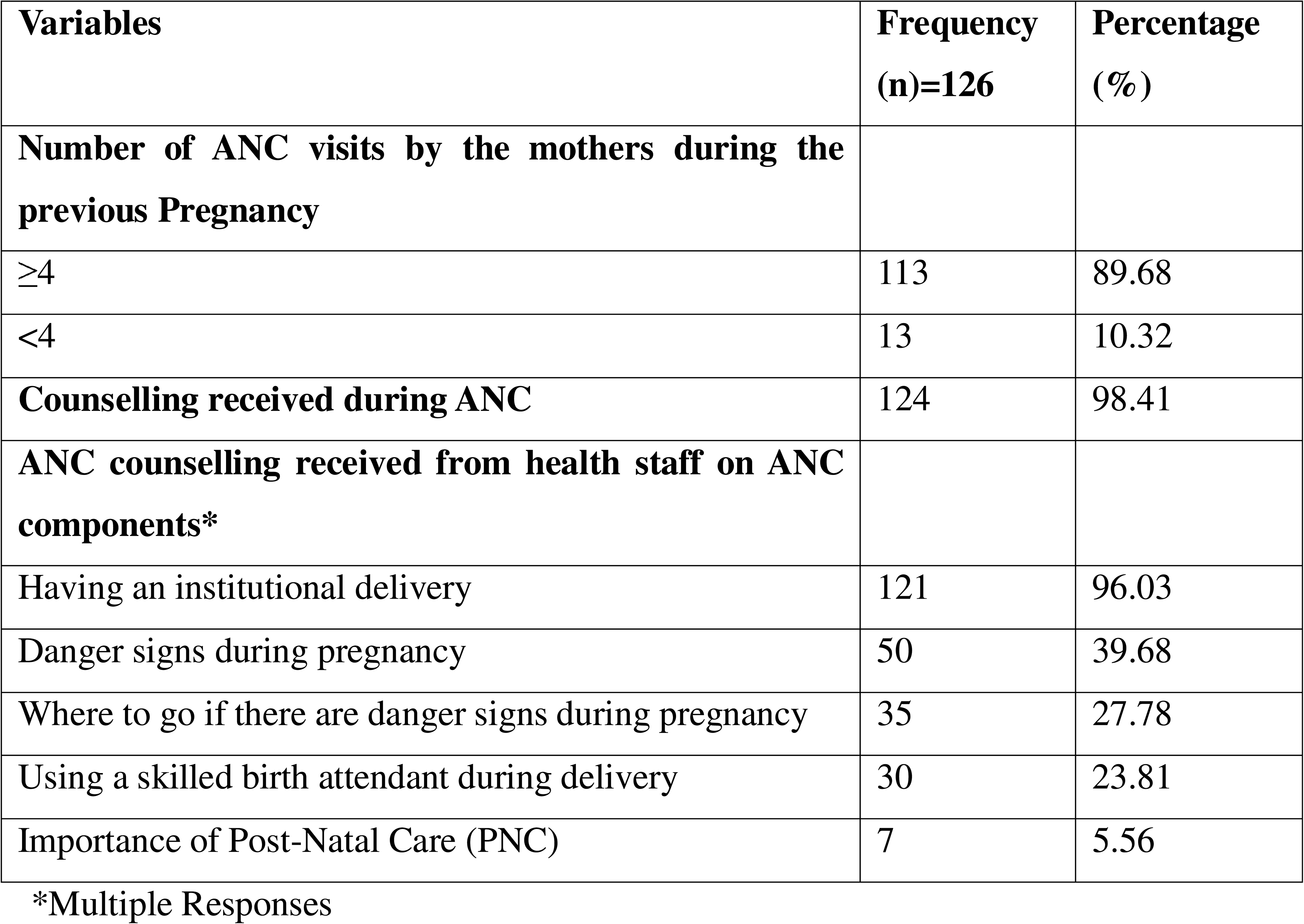
Number of ANC and counseling on ANC during previous pregnancy.

Most (88.09%) of the mothers had information about birth preparedness measures during delivery. Most of the respondents and their family members (86.51%) adopted birth preparedness measures during their last delivery. Among the components of birth preparedness measures, almost all (83.33%) of the mother and their family members saved money for emergency services (Table 3).

**Table 3:** Birth preparedness of mothers during last pregnancy.

| **Variables** | **Frequency**<br>**(n)=126** | **Percentage**<br>**(%)** |
| --- | --- | --- |
| **Mothers had information about birth preparedness during delivery** | 111 | 88.09 |
| **Information on components of birth preparedness Measures\*** |  |  |
| Saving money for emergency services | 110 | 87.30 |
| Arrange transportation beforehand based on local conditions | 65 | 51.58 |
| Identify and contact health facilities and health workers who have a safe delivery kit | 38 | 30.16 |
| Arrange food and clothes | 30 | 23.81 |
| Identify persons who can and are eligible to donate blood | 6 | 4.76 |
| **Mothers and their family members adopted birth preparedness measures** | 109 | 86.51 |
| **Components of birth preparedness measured by the mothers and their family members\*** |  |  |
| Saved money for emergency services | 105 | 83.33 |
| Arranged transportation based on local conditions | 52 | 41.27 |
| Identified and contacted health facilities and health workers having a safe delivery kit | 35 | 27.78 |
| Arranged food and clothes | 22 | 17.46 |
| Identified persons who can and are eligible to donate blood | 1 | 0.79 |

\*Multiple Responses

More than two-fifths (46.82%) of the mothers had information on danger signs of pregnancy. Regarding the components of danger signs during pregnancy, more than one-fourth (37.30%) of the respondent had information that vaginal bleeding was a danger sign during pregnancy and 2.38 % of them knew that convulsion and fits were danger signs during pregnancy (Table 4).

**Table 4:** Information on danger signs during pregnancy among mothers.

| **Variables** | **Frequency**<br>**(n)=126** | **Percentage**<br>**(%)** |
| --- | --- | --- |
| **Respondent had information on the danger signs of pregnancy** | 59 | 46.82 |
| **Components of danger signs during pregnancy\*** |  |  |
| Vaginal bleeding | 47 | 37.30 |
| Severe headache | 25 | 19.84 |
| Severe abdominal pain | 23 | 18.25 |
| Swelling of the hand and face, and blurred vision | 22 | 17.46 |
| Convulsion and fits | 3 | 2.38 |

\*Multiple Responses

More than half (56.35%) of the respondents were first pregnant at ≥20 years. The majority (66.67%) of the respondents had a birth order of more than one. More than one-fourth (29.4%) of the respondents experienced a complication during the most recent pregnancy or childbirth. Most (94.4%) of the respondents had information about the incentive scheme on four ANC and institutional delivery under the “Safe Motherhood Program”. A majority (73.81%) of the respondents were involved in decision-making for the place of delivery during their last pregnancy. More than half (53.97%) of the mothers’ houses were located within less than one hour’s distance from the birthing centre (Table 5).

**Table 5:**
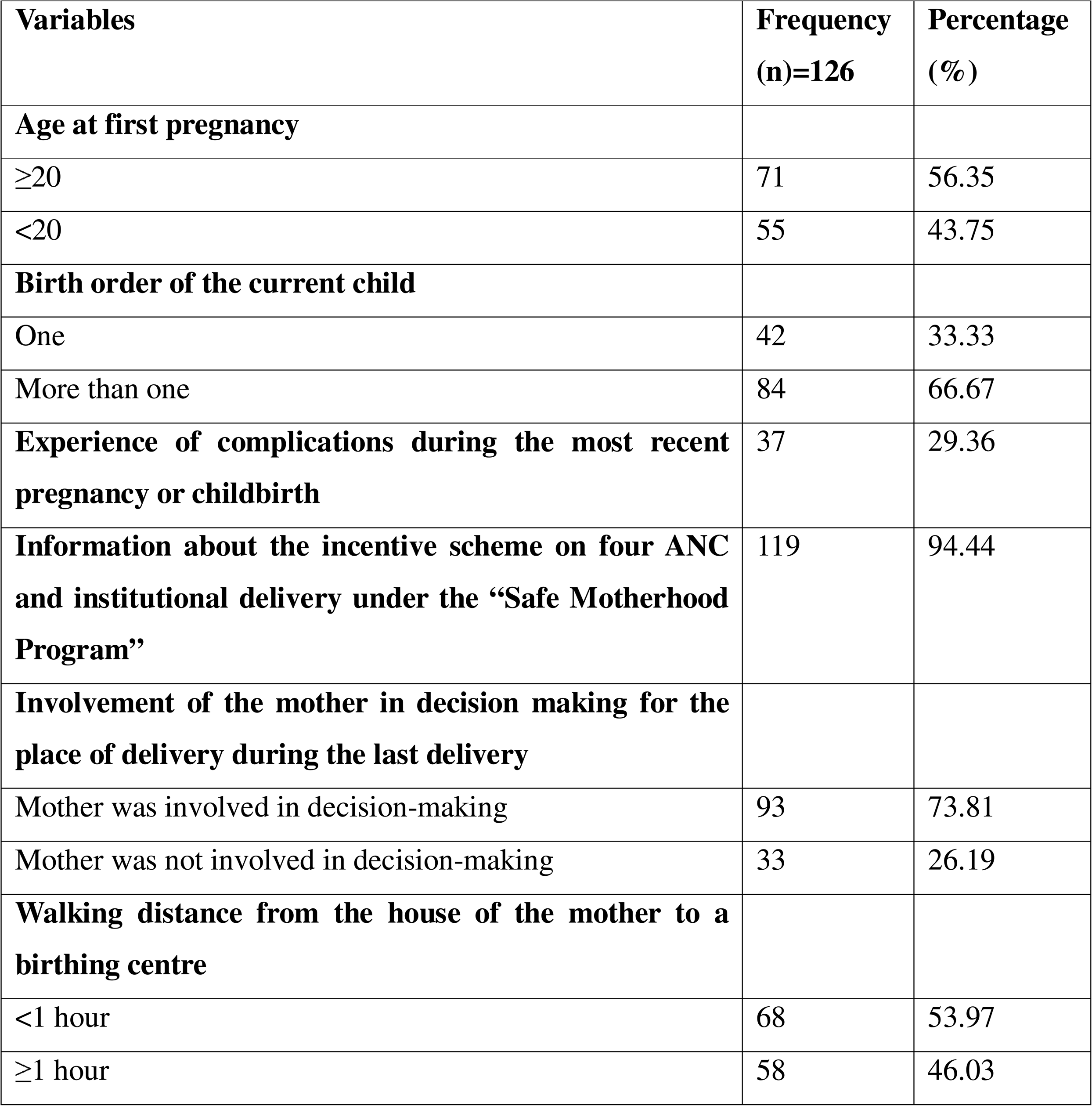
Determinants of maternal health service utilization.

The prevalence of institutional delivery was 90.48%. Most (89.68%) of the mothers were informed by a health worker regarding institutional delivery, and a majority (75.39%) of the mothers were informed by a Female Community Health Volunteer (FCHV) regarding institutional delivery (Table 6).

**Table 6:**
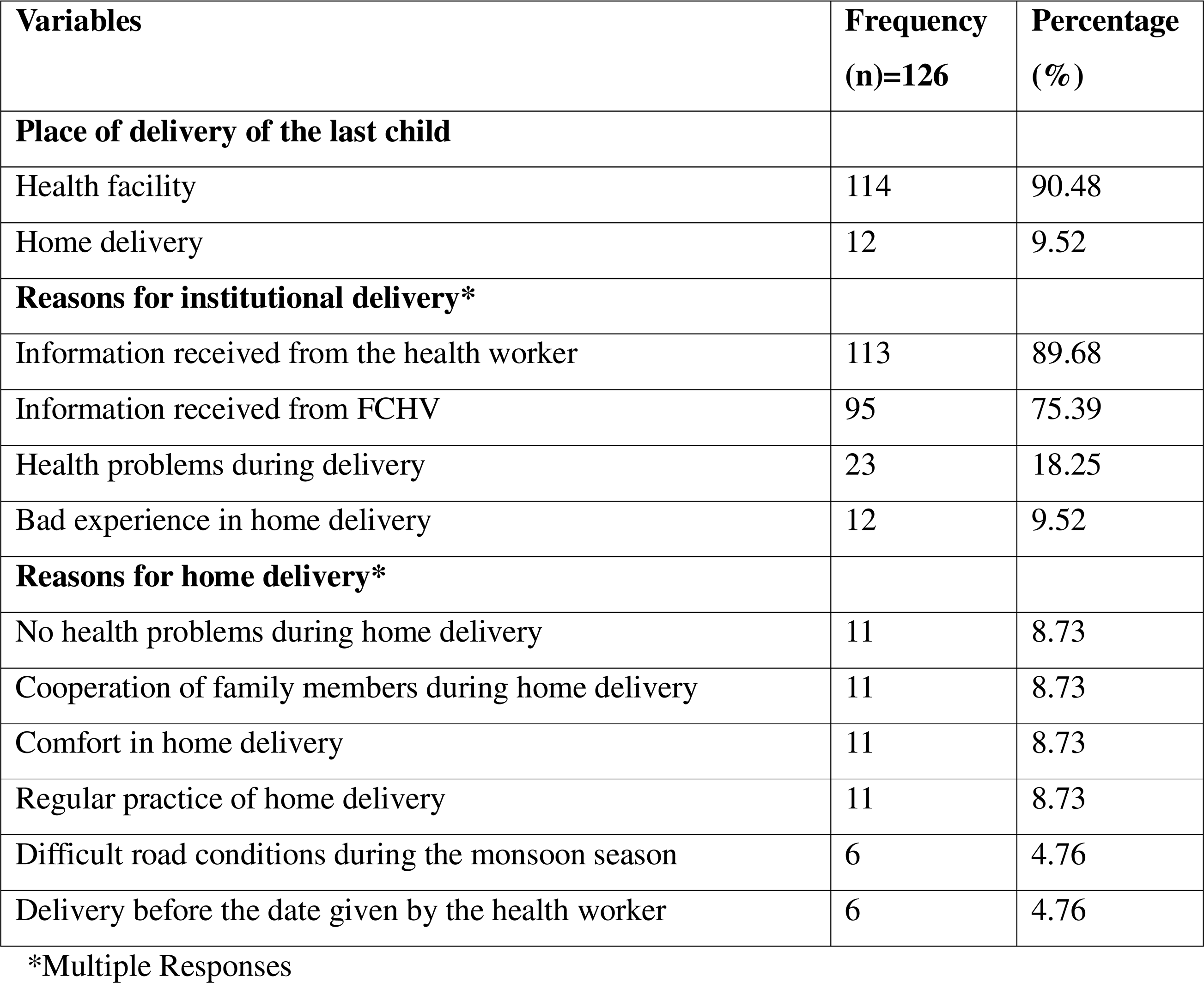
Place of delivery of mother during her last pregnancy and reasons for choice of delivery location.

| Variables | Frequency<br>(n)=126 | Percentage<br>(%) |
| --- | --- | --- |
| **Place of delivery of the last child** |  |  |
| Health facility | 114 | 90.48 |
| Home delivery | 12 | 9.52 |
| **Reasons for institutional delivery\*** |  |  |
| Information received from the health worker | 113 | 89.68 |
| Information received from FCHV | 95 | 75.39 |
| Health problems during delivery | 23 | 18.25 |
| Bad experience in home delivery | 12 | 9.52 |
| **Reasons for home delivery\*** |  |  |
| No health problems during home delivery | 11 | 8.73 |
| Cooperation of family members during home delivery | 11 | 8.73 |
| Comfort in home delivery | 11 | 8.73 |
| Regular practice of home delivery | 11 | 8.73 |
| Difficult road conditions during the monsoon season | 6 | 4.76 |
| Delivery before the date given by the health worker | 6 | 4.76 |

\*Multiple Responses

There was a statistically significant association between the birth order of the current child and institutional delivery, controlling the potential confounding variables, since the p-value from the regression analysis was less than 0.05 (p=0.04). Mothers with more than one child had 0.11 times the odds of having an institutional delivery than first-time mothers (95% CI: 0.01-0.97) (Table 7).

**Table 7:**
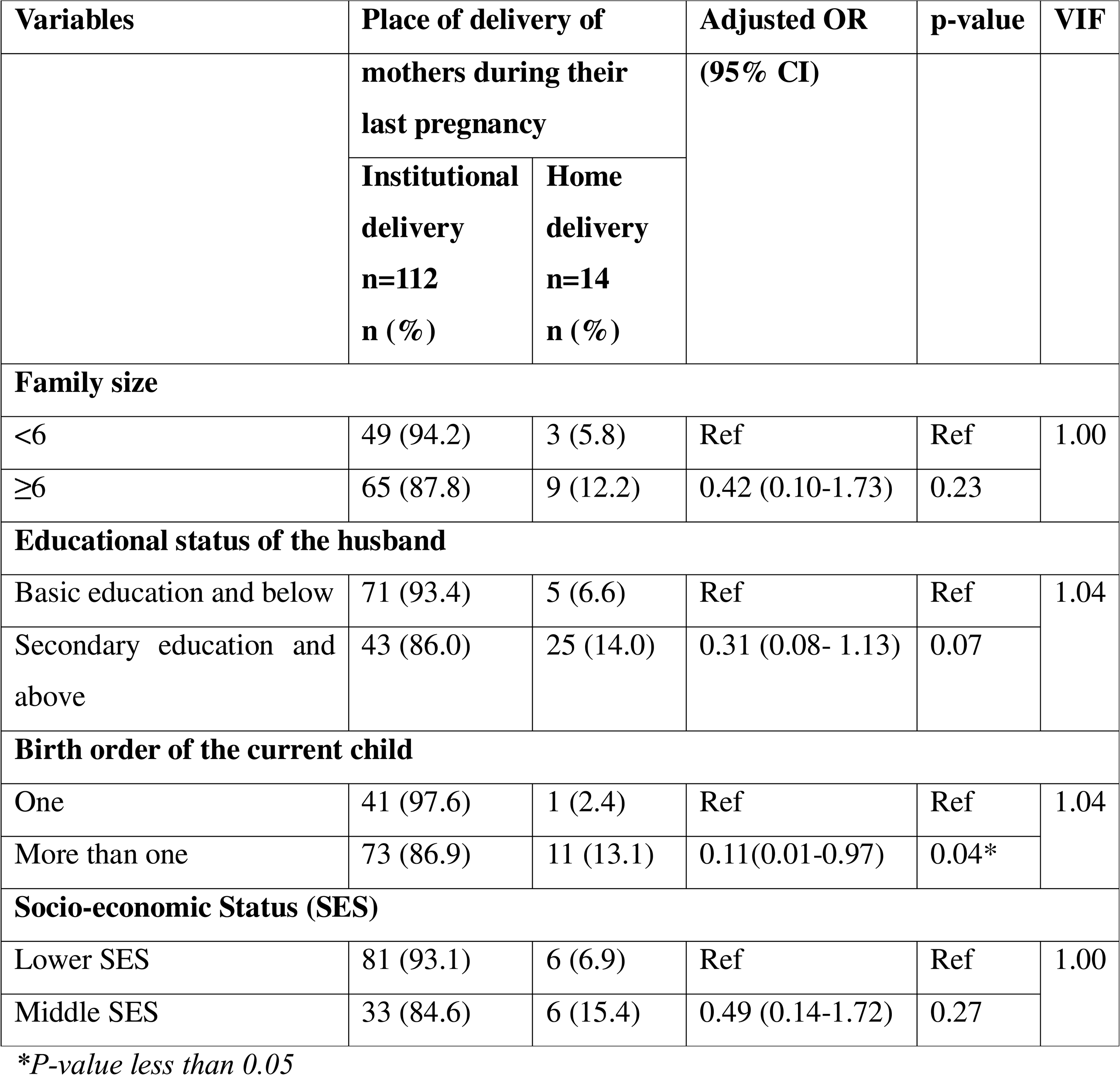
Multivariate model for the association of independent variables and institutional delivery of mothers during their last pregnancy.

## Discussion

This research study found that the prevalence of institutional delivery was 90.48%, which was substantially higher compared to findings from several other research studies in various regions of Nepal. A cross-sectional study conducted by Bhaskar et.al (2018) in Sunsari district, Nepal, reported a significantly lower institutional delivery rate of 55.1% (15). Similarly, a study conducted by Chaurasiya et.al (2019) in Mahottari district, Nepal, found that only 30% of mothers from the Dalit ethnic group had institutional delivery(9). Another study conducted by Pathak et.al (2017) in Chitwan district, Nepal, found that 78.3% of mothers had their delivery in the health facilities(16). It also exceeds the national and provincial institutional delivery prevalence of 79% and 88%, respectively, as per NDHS 2022(17). This discrepancy may be due to regional disparities in healthcare access and socioeconomic factors. The higher rate observed in the present study could also be due to active local government interventions, such as the “Vice-President Delivery House Visit Program” and the provision of an additional Rs. 1000 incentive under the “Safe Motherhood Program” from the Tamankhola rural municipality. These financial and social incentives, budgeted by the Tamankhola Rural Municipality, likely played a critical role in encouraging more women to have institutional deliveries(18).

The research study found that mothers with more than one child had 0.11 times the odds of having an institutional delivery than first-time mothers (95% CI: 0.01-0.97). These findings are consistent with a study conducted by Joshi et.al (2016) in Mugu district, Nepal which found that primiparous mothers had higher odds of institutional delivery [adjusted OR (aOR) 2.3, 95%CI 1.1–4.8] than multiparous mothers (19) and further analysis of NDHS 2016 by Parajuli et.al (2023) found that primiparous mothers had higher odds of institutional delivery [aOR: 2.79; 95% CI: 2.26-3.46] (20). The underlying reasons may include service satisfaction during institutional delivery by primiparous mothers, geographical and financial constraints to reach birthing centers, and underestimation of home delivery risks by multiparous women.

### Strengths and limitations

The tools of the research study were formulated from an extensive literature review and validated by a research supervisor and faculty of the Central Department of Public Health, IoM, to ensure content validity. Face validity was ensured by conducting a pre-testing of the research tool. Multivariate logistic regression was used for quantitative statistical analysis, controlling the potential confounding variables, and VIF was also calculated to assess multicollinearity.

The health facility and health service provider related factors were not taken into account as factors influencing the utilization of institutional delivery services by mothers.

## Conclusions

The study found that the prevalence of institutional delivery was 90.48%, and primiparous mothers had higher odds of institutional delivery.

Majority of the mothers reported attending ANC visits as per protocol during their last childbirth, and almost all of them received counseling on institutional delivery. However, the quality and comprehensiveness of ANC counseling remain inadequate, as only 5.56% were counseled on the importance of PNC visits. Key components of birth preparedness were frequently overlooked. Knowledge of pregnancy danger signs was also limited.

Though the prevalence of institutional delivery was higher than provincial and national prevalence, 9.52% of the mothers still delivered at home. Poor road conditions, particularly during the monsoon season, were identified as a key barrier to facility-based delivery.

This highlights the importance of improving the quality of health education and removing structural barriers to care. The health section chief of the Tamankhola rural municipality should strengthen its coordination with health staff and FCHVs to ensure comprehensive ANC counseling, increase awareness about danger signs, and promote all aspects of birth preparedness. Additionally, the health section chief should coordinate with the Department of Local Infrastructure (DoLI) for improving road infrastructure to ensure safe and timely access to health facilities, particularly during adverse weather conditions, to further advance maternal and newborn health outcomes in Tamankhola rural municipality.

## Data Availability

All data produed in the present study are available upon reasonable request to the authors.

## Acknowledgments

The study acknowledges the Health Section Chief of Tamankhola Rural Municipality, mothers who participated in this research study, and all the supporting hands for their invaluable contributions to this research journey.

---

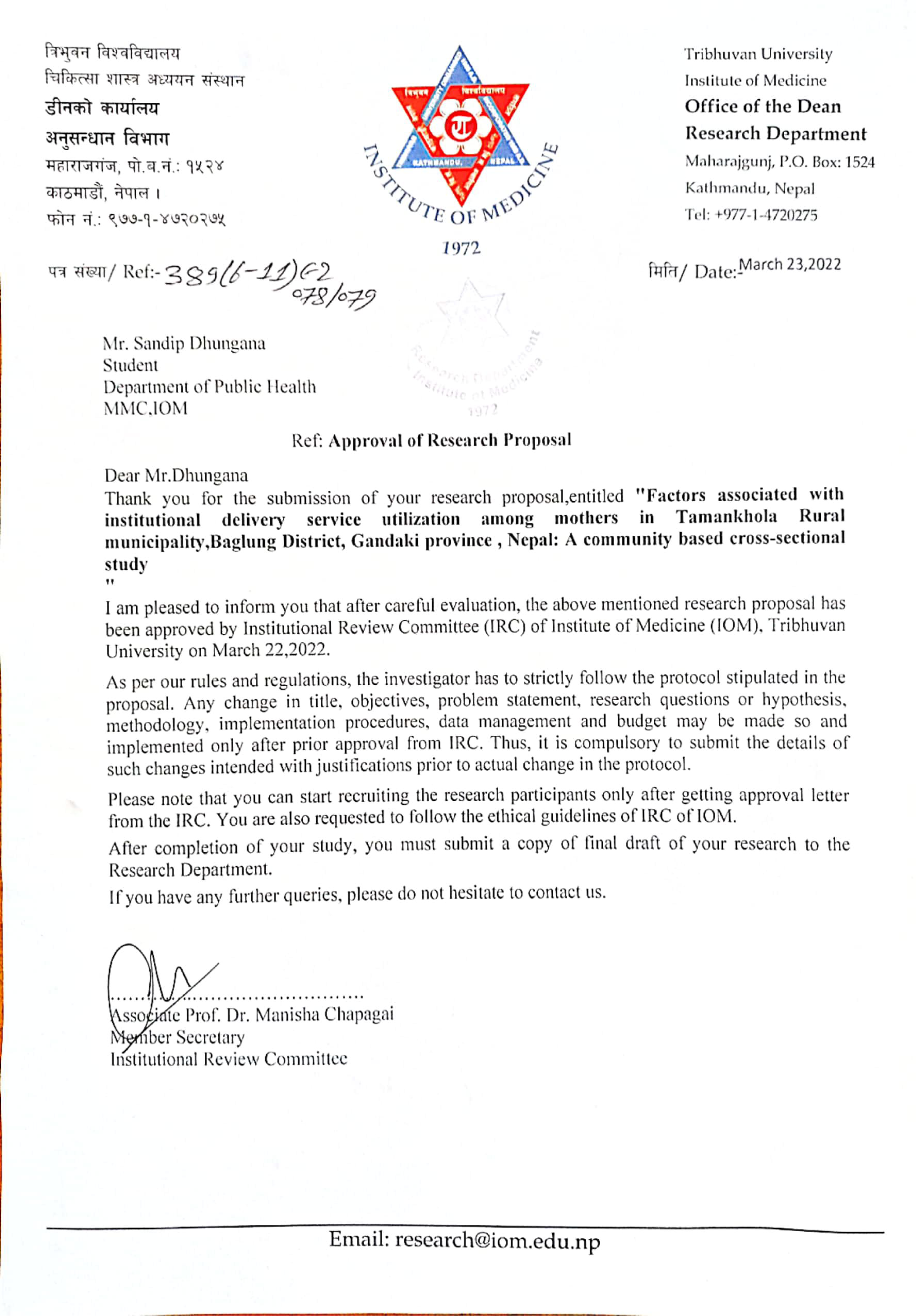

## References

1. Sagun’s blog. Health Contents - Constitution of Nepal [Internet]. 2018 [cited 2025 May 8]. Available from: https://publichealthupdate.com/health-contents-constitution-of-nepal/

2. Center For Reproductive Rights. Safe Motherhood and Reproductive Health Rights Act, 2018 Unofficial Translation. 2018 Sep 18 [cited 2025 May 8]; Available from: https://nepal.unfpa.org/sites/default/files/pub-pdf/The%20Right%20to%20Safe%20Motherhood%20and%20Reproductive%20Health%20Act%202075.pdf

3. Department of Health Services. Public Health Update. 2024 [cited 2024 Jun 11]. DoHS_AHR Book - Final 26 Feb 2024_Public Health Update - Google Drive. Available from: https://drive.google.com/file/d/12cvDnPC5ecuGHIXfq4y77pAtKxC6UP47/view

4. Ministry of Health and Population G of N. National Policy on Skilled Birth Attendants. 2006 Jul [cited 2025 May 8]; Available from: https://giwmscdntwo.gov.np/media/pdf_upload/Safe%20Motherhood%20and%20SBA%20Policy_94rbjzq.pdf

5. New ERA. Nepal Demographic and Health Survey 2022 Key Indicators Report Ministry of Health and Population New ERA Ministry of Health and Population. 2023 Jun [cited 2025 Apr 12]; Available from: http://www.mohp.org.np;

6. USAID+New ERA+ Ministry of Health and Population. 2016 Demographic and Health Survey Key Findings Nepal. 2017 [cited 2025 May 8]; Available from: https://nepal.unfpa.org/sites/default/files/pub-pdf/NDHS%202016%20key%20findings.pdf

7. Tamankhola Rural Municipality. Tamankhola Rural Municipality, Bagmati Province, Nepal [Internet]. 2025 [cited 2025 May 8]. Available from: https://tamankholamun.gov.np/

8. National Statistical Office G of N. A report on maternal mortality, National Population and Housing Census 2021. 2021;

9. Chaurasiya SP, Pravana NK, Khanal V, Giri D. Two thirds of the most disadvantaged Dalit population of Nepal still do not deliver in health facilities despite impressive success in maternal health. PLoS One [Internet]. 2019 Jun 1 [cited 2025 May 14];14(6):e0217337. Available from: https://journals.plos.org/plosone/article?id=10.1371/journal.pone.0217337

10. Tamankhola Rural Municipality. Annual Development Plan. 2025;

11. Shah R, Rehfuess EA, Maskey MK, Fischer R, Bhandari PB, Delius M. Factors affecting institutional delivery in rural Chitwan district of Nepal: A community-based cross-sectional study. BMC Pregnancy Childbirth [Internet]. 2015 Feb 13 [cited 2025 May 8];15(1):1–14. Available from: https://bmcpregnancychildbirth.biomedcentral.com/articles/10.1186/s12884-015-0454-y

12. Sürücü L, Maslakçi A. VALIDITY AND RELIABILITY IN QUANTITATIVE RESEARCH. Business & Management Studies: An International Journal. 2020 Sep 25;8(3):2694–726.

13. Razali MNWBY. (PDF) Power Comparisons of Shapiro-Wilk, Kolmogorov-Smirnov, Lilliefors and Anderson-Darling Tests [Internet]. 2011 [cited 2025 Apr 13]. Available from: https://www.researchgate.net/publication/267205556_Power_Comparisons_of_Shapiro-Wilk_Kolmogorov-Smirnov_Lilliefors_and_Anderson-Darling_Tests

14. Study/Books_Need2Read/David W. Hosmer - Applied Logistic Regression - 3rd Edition.pdf at master · Drxan/Study · GitHub [Internet]. [cited 2024 Jul 20]. Available from: https://github.com/Drxan/Study/blob/master/Books_Need2Read/David%20W.%20Hosmer%20-%20Applied%20Logistic%20Regression%20-%203rd%20Edition.pdf

15. Author C, Kumar Bhaskar R, Ravi Kumar B, Krishna Kumar D. Determinants of Utilization of Institutional Delivery Services in East Nepal: A Community-Based Cross-Sectional Study. Med Phoenix [Internet]. 2018 Aug 15 [cited 2025 May 8];3(1):6–15. Available from: https://www.nepjol.info/index.php/medphoenix/article/view/20755

16. Pathak P, Shrestha S, Devkota R, Thapa B. Factors Associated with the Utilization of Institutional Delivery Service among Mothers. J Nepal Health Res Counc [Internet]. 2017 [cited 2025 May 14];15(3):228–34. Available from: https://www.nepjol.info/index.php/JNHRC/article/view/18845

17. Ministry of Health and Population+New ERA. Nepal Demographic and Health Survey 2022 Key Indicators Report. 2022 [cited 2024 Jun 11]; Available from: http://www.mohp.org.np;

18. Tamankhola Rural Municipality. Budget and program for Fiscal Year 2079/2080 B.S. 2022 [cited 2025 May 15]; Available from: https://tamankholamun.gov.np/sites/tamankholamun.gov.np/files/2079.080.pdf

19. Joshi D, Baral SC, Giri S, Kumar AM V. Universal institutional delivery among mothers in a remote mountain district of Nepal: what are the challenges? Public Health Action [Internet]. 2016 Dec 28 [cited 2025 May 14];6(4):267. Available from: https://pmc.ncbi.nlm.nih.gov/articles/PMC5176053/

20. Parajuli A, K.C. J, Magar EB, Chaudhary R, Gupta M, Marahatta D. Predictors of Institutional Delivery in Nepal: Analyzing Nepal Demographic and Health Survey 2016. Journal of Multidisciplinary Research Advancements. 2023 Dec 31;1(2):104–13.

